# Effect of Prophylactic Care Bundle on Acute Kidney Injury Following Coronary Artery Bypass Grafting

**DOI:** 10.1101/2025.11.26.25341121

**Authors:** Heng Zhang, Jiawei Han, Hongyan Zhou, Fangzhou Li, Lili Liu, Yan Zhao, Ge Gao, Juan Du, Jianfang Cai, Sheng Liu

## Abstract

**BACKGROUND:** Acute kidney injury (AKI) following cardiac surgery is common, yet no consistently effective preventive strategy has been established. Therefore, the aim of this study was to assess the effectiveness of perioperative care bundle in preventing AKI following cardiac surgery.

**METHODS:** The study included consecutive low-risk and high-risk patients who underwent coronary artery bypass grafting (CABG) at China National Center for Cardiovascular Diseases from January 1, 2020, to September 30, 2022. The intervention consisted of a structured care bundle based on Kidney Disease: Improving Global Outcomes guidelines, including close hemodynamic monitoring, goal-directed fluid management, avoidance of nephrotoxins, temporary discontinuation of angiotensin-converting-enzyme inhibitors/angiotensin II receptor blockers for 48 h, serial monitoring of serum creatinine and urine output, and prevention of hyperglycemia. The primary outcome was AKI within 7 days post-surgery. The secondary outcome was in-hospital major adverse cardiac and kidney events (composite of death, renal replacement therapy, >200% creatinine increase, myocardial infarction, or stroke).

**RESULTS:** Of 8,603 patients (2,462 receiving bundle care; 6,141 usual care), 2,416 propensity-matched pairs were analyzed. Mean age was 60.2 years (bundle: standard deviation [SD] 8.9; usual care: SD 9.0); 16.5% vs. 16.3% were women. AKI risk was significantly lower with bundle care vs. usual care (53.1% vs. 59.9%; mean difference, –6.8% [95% confidence interval (CI), –9.6% to –0.4%]; relative risk [RR], 0.89 [95% CI, 0.84 to 0.93]; *P* < .001). Bundle care also reduced major adverse cardiac and kidney events (4.7% vs. 6.3%; mean difference, −1.6% [95% CI, −2.9% to −0.3%]; RR, 0.75 [95% CI, 0.59 to 0.95]; *P* = .02). Benefits were more pronounced in high-risk patients (for AKI, RR, 0.89 [95% CI, 0.84 to 0.95]; *P* < .001; for major adverse cardiac and kidney events, RR, 0.74 [95% CI, 0.55 to 1.00]; *P* = .04). No statistically significant difference in all-cause mortality was observed.

**CONCLUSIONS:** Among patients undergoing CABG, perioperative care bundle strategy, compared with standard care, was significantly associated with a lower risk of AKI and in-hospital major adverse cardiac and kidney events. Randomized clinical trials are warranted to confirm these findings.

## Introduction

The prevalence of acute kidney injury (AKI) among patients with cardiovascular disease continues to rise, particularly among those undergoing cardiac surgeries, leading to increased mortality and substantial healthcare costs.^1,2^ Effective management of AKI requires a multifaceted approach encompassing early diagnosis, monitoring of renal function, and implementation of preventive and therapeutic interventions.^3^ However, no universally accepted therapy has consistently demonstrated improved renal function recovery or clinical outcomes in patients with AKI.^4,5^

Conventional AKI management focuses on optimizing volume status and hemodynamics, avoiding nephrotoxic drugs, and preventing hyperglycemia in high-risk patients.^6,7^ Therefore, care bundles have been proposed to improve the quality of care and outcome of patients with AKI.^8^ An effective care bundle typically includes a concise set of evidence-based interventions recommended by clinical guidelines and widely endorsed by local stakeholders.^9,10^ Although several AKI care bundles have been implemented with variable improvement in clinical care, particularly when combined with educational measures and electronic alert systems, large, adequately powered studies are warranted to investigate their impact on mortality and renal outcomes.^11–13^

This study aimed to evaluate the association of AKI care bundle adherence and postoperative cardiovascular and renal outcomes in patients undergoing coronary artery bypass grafting (CABG).

## Methods

### Study Design and Data

The observational study included consecutive patients who underwent CABG at China National Center for Cardiovascular Diseases from January 1, 2020, to September 30, 2022. Data were obtained from the institutional medical records linkage system, which comprehensively captures hospital admissions, procedures, and outcomes according to definitions of the Society of Thoracic Surgeons National Adult Cardiac Database. Efforts were made to ensure the accuracy and completeness of this data have been described previously (eMethod 1 in Supplement).^14,15^ The study was approved by the Fuwai Hospital Institutional Review Board, which granted a waiver of informed consent owing to the retrospective nature and minimal risk nature to participants.

### Study Population and Intervention

Adult patients (aged ≥18 years) undergoing CABG with or without concomitant valvular heart surgery or ascending aortic surgery at Fuwai Hospital were eligible. Exclusion criteria included ongoing dialysis, prior nephrectomy, or kidney transplant within the last 12 months. Considering the potential influence of AKI risk before CABG, this study assessed the risk of developing AKI and clinical outcomes in two separate cohorts: patients with low risk and patients with high risk of AKI. Patients were recorded as high risk according to the presence of at least one of the following risk factors: age >70 years, estimated glomerular filtration (eGFR) <60 mL/min/1.73m^2^, diabetes, congestive heart failure, and those who received concomitant cardiac surgery other than CABG.^16^

General anesthesia was induced with propofol and maintained using sevoflurane and propofol. All CABG procedures were performed with standard bypass techniques (eMethod 2 in Supplement). The intervention consisted of structured care bundle according to Kidney Disease Improving Global Outcomes (KDIGO) practice guidelines. Components included serial serum creatinine and urine output monitoring, close hemodynamic monitoring with goal directed volume resuscitation, avoidance of nephrotoxic substances, temporary discontinuation of angiotensin-converting-enzyme inhibitors (ACEIs) and angiotensin II receptor blockers (ARBs) for 48 h, and prevention of hyperglycaemia (eMethod 3 in Supplement).^8,17^ The use of diuretics, initiation of renal replacement therapy, and ventilation management were determined by the treating physician according to current guidelines. Complications during intensive care unit stay were also recorded, including additional emergent surgery, major bleeding events, transfusion requirements, and arrhythmias.

### Outcomes

The primary outcome was occurrence of AKI (defined by serum creatinine-based and urine output criteria from the KDIGO guidelines, eMethod 4 in Supplement) within the 7 days postoperatively.^10^ Baseline serum creatinine level was determined from the last preoperative measurement. Postoperative serum creatinine was measured at least once daily for the first 7 days after CABG, and urine output monitored up to postoperative day 4 in most patients, because urinary catheters are routinely removed on discharge from the intensive care unit. The secondary outcome was the occurrence of in-hospital major adverse cardiac and kidney events, a composite of death, renal replacement therapy, >200% sustained increase of creatinine from baseline, myocardial infarction and stroke after CABG procedure.^18^ Myocardial infarction and stroke were identified using cardiovascular-related diagnostic codes, including the International Classification of Diseases, and the American Heart Association definitions (eMethod 5 in Supplement).^19^

Subgroup analysis of the primary and secondary outcomes was performed for demographics (age and sex), comorbidities, baseline renal and cardiac strata, and surgical categories. Post hoc analyses included several exploratory endpoints including major surgical complications, length of intensive care unit, and hospital stay. Risk factors for AKI were also analyzed.

### Statistical Analysis

Categorical variables were expressed as frequencies and percentages and continuous variables were expressed as the mean and standard deviation (SD) or median and interquartile range, depending on the distribution. We used the Pearson χ^2^ test and the Wilcoxon rank sum test to compare the randomized treatment groups regarding categorical and continuous outcomes, respectively.

Propensity score matching was used to adjust for baseline differences between bundle and usual care groups. Propensity scores were estimated via logistic regression incorporating sociodemographics, procedure-related characteristics, medical history, concurrent medication use, surgery year.^20^ Covariate balance between the groups was assessed by calculating standardized differences for which a difference of <0.10 was considered to indicate good balance.^21^ The mean differences are the estimated differences in the geometric means between the bundle and usual care groups.

Laboratory data were used to assess values for serum creatinine and hemoglobin. These laboratory data were not included in the model to calculate the propensity score, but the values and the proportion of missing values were balanced after matching on all other patient characteristics. Balance of baseline characteristics was also assessed in patients with low and high risk of AKI, separately, because the comparisons between patients treated with and without bundle care were stratified by the presence of AKI risk. The difference in the probability of outcome events in the matched cohorts was evaluated by the McNemar test and paired *t* tests when appropriate. Relative risk (RR) was estimated as the ratio of the probability of the outcome event in patients treated using the bundle care compared with patients treated using the usual care. The 95% confidence interval (CI) was constructed using methods that accounted for the matched nature of the cohorts.

Prespecified subgroup analyses were stratified by age, sex, baseline eGFR, diabetes, left ventricle function impairment, CABG type, use of cardiopulmonary bypass, and AKI risk. Heterogeneity was estimated using χ^2^ test for heterogeneity and the *I*^2^ statistic. Whether the RRs were the same across the subgroups was tested by the significance of the interaction terms. To assess factors associated with AKI, independent logistic regression analyses were performed for each factor. Whenever mean, median, or percentage difference were reported, the 95% CI was computed with the t test, bootstrapping, and Wilson method with continuity correction, respectively.

Analyses of secondary and post hoc endpoints were considered exploratory owing to potential type I error from multiple comparisons. All statistical analyses were performed using R version 3.3.1 (R Foundation), with two-sided *P* < .05 considered statistically significant.

## Results

### Baseline Characteristics

Of 10,883 patients screened in Fuwai Cardiovascular Surgery Registry between January 2020 and September 2022, 2,227 were excluded because of missing laboratory or hemodynamic data, 8 because of preoperative dialysis, and 42 because of prior nephrectomy or kidney transplant. Among the 8,603 eligible patients, the mean (SD) age was 61.2 (8.8) years, 1,844 (21.4%) were women, 3,940 (45.8%) had diabetes, 173 (2.0%) had preexisting chronic renal failure, and 2,313 (26.9%) underwent concomitant cardiac surgery. Table 1 shows patient demographics at baseline before and after propensity score matching. Those who received care bundle strategy were less likely to have diagnostic codes for hypertension, diabetes, previous stroke, and congestive heart failure; and more likely to have isolated CABG. After matching, 2,416 pairs of patients remained in the study population and the two cohorts were well matched. Of these patients, 2698/4832 (55.8%) had a high risk of developing AKI; baseline characteristics stratified by whether patients had a high or low risk of AKI are reported in eTable 1 in Supplement.

**Table 1.**
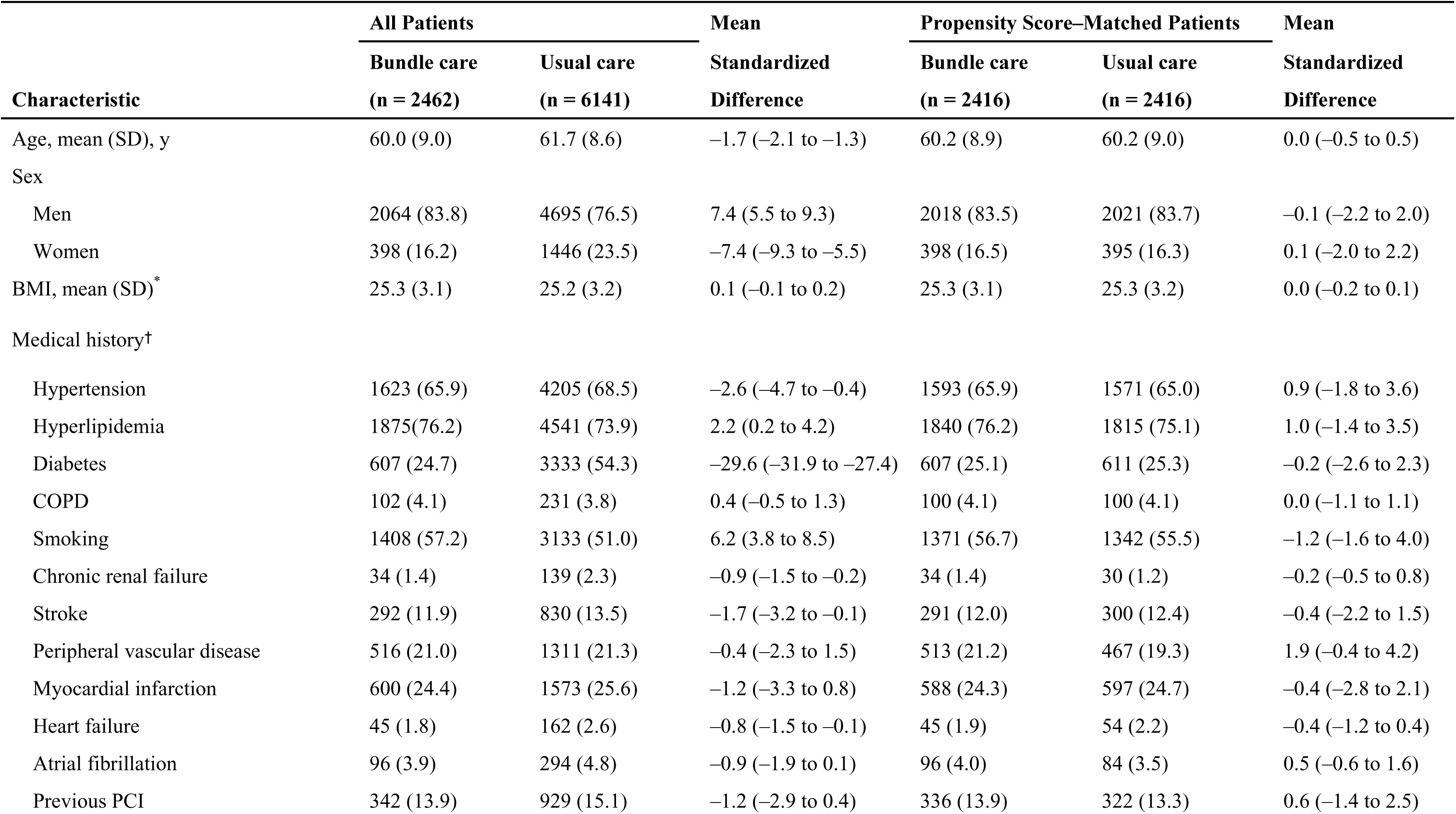
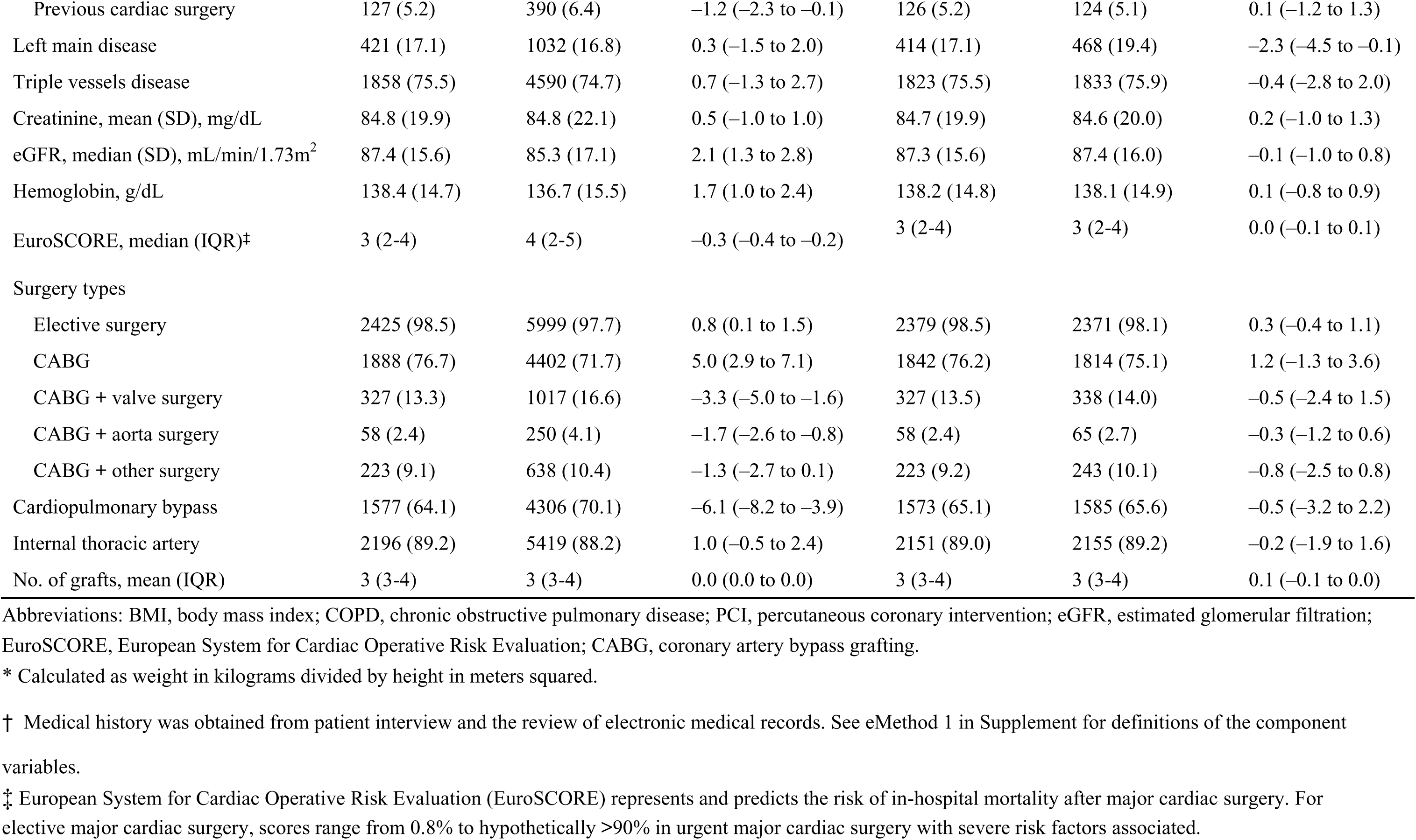
Baseline Demographic, Clinical, and Procedural Characteristics.

### Analyses of AKI

The primary outcome of AKI occurred in 1,282 of 2,416 (53.1%) in the bundle care group compared with 1,446 of 2,416 (59.9%) in the usual care group with a risk difference of –6.8% percentage points (95% CI, −9.6% to –0.4%; RR, 0.89; 95% CI, 0.84 to 0.93; *P* < .001) (Table 2). In patients with low risk, the incidence of AKI was 48.3% in patients with bundle care and 54.4% in patients with bundle care (mean difference, –6.1% [95% CI, –10.1% to –1.8%]; RR, 0.89; 95% CI, 0.82 to 0.97; *P* = .01). However, the risks of stage III AKI were not significantly different between groups. In patients with high-risk, a statistically significant decreased risk of AKI in almost all stages was observed for the bundle care group compared with the usual care group. Multiple logistic regression analysis showed that patients with high risk, age ≥ 70 years, hypertension, diabetes, chronic renal failure, and eGFR < 60 mL/min/1.73m^2^ were independently associated with the incidence of AKI (eFigure 2 in Supplement).

**Table 2.**
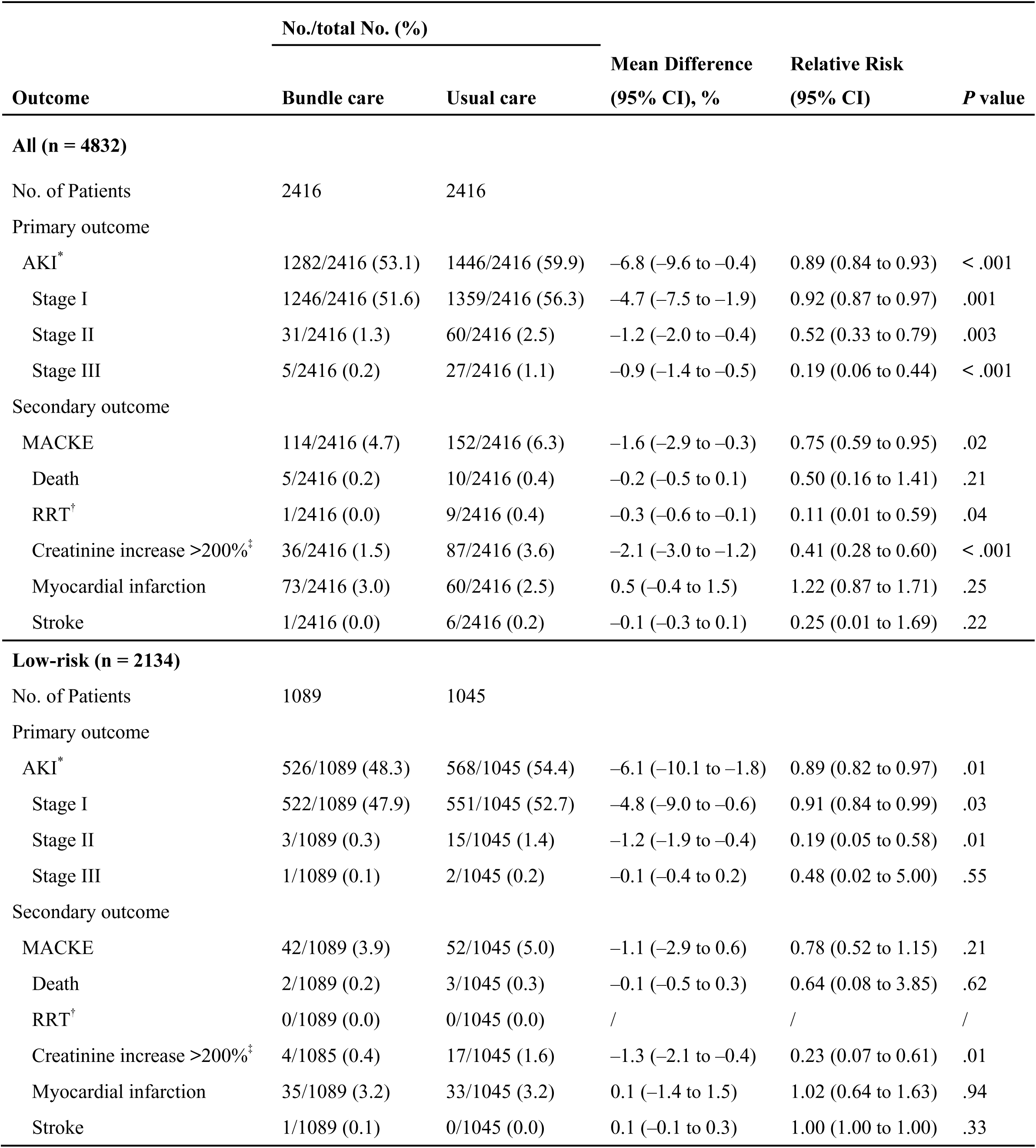
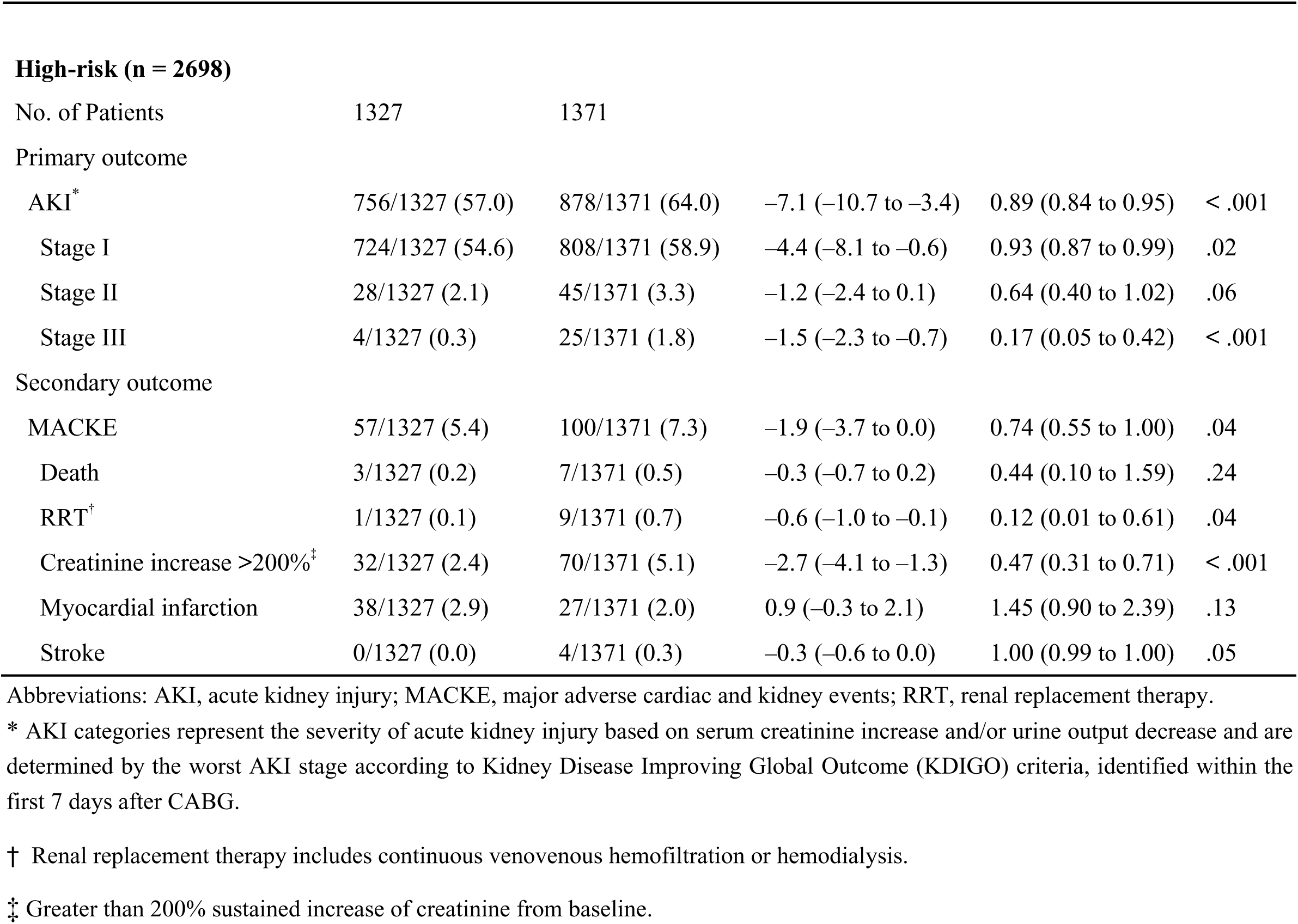
Effect of the Care Bundle on the Primary and Secondary Outcomes.

|  | No./total No. (%) |  |  |  |  |
| --- | --- | --- | --- | --- | --- |
| Outcome | Bundle care | Usual care | Mean Difference<br>(95% CI), % | Relative Risk<br>(95% CI) | P value |
| All (n = 4832) |  |  |  |  |  |
| No. of Patients | 2416 | 2416 |  |  |  |
| Primary outcome |  |  |  |  |  |
| AKI* | 1282/2416 (53.1) | 1446/2416 (59.9) | −6.8 (−9.6 to −0.4) | 0.89 (0.84 to 0.93) | < .001 |
| Stage I | 1246/2416 (51.6) | 1359/2416 (56.3) | −4.7 (−7.5 to −1.9) | 0.92 (0.87 to 0.97) | .001 |
| Stage II | 31/2416 (1.3) | 60/2416 (2.5) | −1.2 (−2.0 to −0.4) | 0.52 (0.33 to 0.79) | .003 |
| Stage III | 5/2416 (0.2) | 27/2416 (1.1) | −0.9 (−1.4 to −0.5) | 0.19 (0.06 to 0.44) | < .001 |
| Secondary outcome |  |  |  |  |  |
| MACKE | 114/2416 (4.7) | 152/2416 (6.3) | −1.6 (−2.9 to −0.3) | 0.75 (0.59 to 0.95) | .02 |
| Death | 5/2416 (0.2) | 10/2416 (0.4) | −0.2 (−0.5 to 0.1) | 0.50 (0.16 to 1.41) | .21 |
| RRT† | 1/2416 (0.0) | 9/2416 (0.4) | −0.3 (−0.6 to −0.1) | 0.11 (0.01 to 0.59) | .04 |
| Creatinine increase >200%‡ | 36/2416 (1.5) | 87/2416 (3.6) | −2.1 (−3.0 to −1.2) | 0.41 (0.28 to 0.60) | < .001 |
| Myocardial infarction | 73/2416 (3.0) | 60/2416 (2.5) | 0.5 (−0.4 to 1.5) | 1.22 (0.87 to 1.71) | .25 |
| Stroke | 1/2416 (0.0) | 6/2416 (0.2) | −0.1 (−0.3 to 0.1) | 0.25 (0.01 to 1.69) | .22 |
| Low-risk (n = 2134) |  |  |  |  |  |
| No. of Patients | 1089 | 1045 |  |  |  |
| Primary outcome |  |  |  |  |  |
| AKI* | 526/1089 (48.3) | 568/1045 (54.4) | −6.1 (−10.1 to −1.8) | 0.89 (0.82 to 0.97) | .01 |
| Stage I | 522/1089 (47.9) | 551/1045 (52.7) | −4.8 (−9.0 to −0.6) | 0.91 (0.84 to 0.99) | .03 |
| Stage II | 3/1089 (0.3) | 15/1045 (1.4) | −1.2 (−1.9 to −0.4) | 0.19 (0.05 to 0.58) | .01 |
| Stage III | 1/1089 (0.1) | 2/1045 (0.2) | −0.1 (−0.4 to 0.2) | 0.48 (0.02 to 5.00) | .55 |
| Secondary outcome |  |  |  |  |  |
| MACKE | 42/1089 (3.9) | 52/1045 (5.0) | −1.1 (−2.9 to 0.6) | 0.78 (0.52 to 1.15) | .21 |
| Death | 2/1089 (0.2) | 3/1045 (0.3) | −0.1 (−0.5 to 0.3) | 0.64 (0.08 to 3.85) | .62 |
| RRT† | 0/1089 (0.0) | 0/1045 (0.0) | / | / | / |
| Creatinine increase >200%‡ | 4/1085 (0.4) | 17/1045 (1.6) | −1.3 (−2.1 to −0.4) | 0.23 (0.07 to 0.61) | .01 |
| Myocardial infarction | 35/1089 (3.2) | 33/1045 (3.2) | 0.1 (−1.4 to 1.5) | 1.02 (0.64 to 1.63) | .94 |
| Stroke | 1/1089 (0.1) | 0/1045 (0.0) | 0.1 (−0.1 to 0.3) | 1.00 (1.00 to 1.00) | .33 |
| **High-risk (n = 2698)** |  |  |  |  |  |
| No. of Patients | 1327 | 1371 |  |  |  |
| Primary outcome |  |  |  |  |  |
| AKI* | 756/1327 (57.0) | 878/1371 (64.0) | −7.1 (−10.7 to −3.4) | 0.89 (0.84 to 0.95) | < .001 |
| Stage I | 724/1327 (54.6) | 808/1371 (58.9) | −4.4 (−8.1 to −0.6) | 0.93 (0.87 to 0.99) | .02 |
| Stage II | 28/1327 (2.1) | 45/1371 (3.3) | −1.2 (−2.4 to 0.1) | 0.64 (0.40 to 1.02) | .06 |
| Stage III | 4/1327 (0.3) | 25/1371 (1.8) | −1.5 (−2.3 to −0.7) | 0.17 (0.05 to 0.42) | < .001 |
| Secondary outcome |  |  |  |  |  |
| MACKE | 57/1327 (5.4) | 100/1371 (7.3) | −1.9 (−3.7 to 0.0) | 0.74 (0.55 to 1.00) | .04 |
| Death | 3/1327 (0.2) | 7/1371 (0.5) | −0.3 (−0.7 to 0.2) | 0.44 (0.10 to 1.59) | .24 |
| RRT† | 1/1327 (0.1) | 9/1371 (0.7) | −0.6 (−1.0 to −0.1) | 0.12 (0.01 to 0.61) | .04 |
| Creatinine increase >200%‡ | 32/1327 (2.4) | 70/1371 (5.1) | −2.7 (−4.1 to −1.3) | 0.47 (0.31 to 0.71) | < .001 |
| Myocardial infarction | 38/1327 (2.9) | 27/1371 (2.0) | 0.9 (−0.3 to 2.1) | 1.45 (0.90 to 2.39) | .13 |
| Stroke | 0/1327 (0.0) | 4/1371 (0.3) | −0.3 (−0.6 to 0.0) | 1.00 (0.99 to 1.00) | .05 |

Abbreviations: AKI, acute kidney injury; MACKE, major adverse cardiac and kidney events; RRT, renal replacement therapy.
\* AKI categories represent the severity of acute kidney injury based on serum creatinine increase and/or urine output decrease and are determined by the worst AKI stage according to Kidney Disease Improving Global Outcome (KDIGO) criteria, identified within the first 7 days after CABG.
† Renal replacement therapy includes continuous venovenous hemofiltration or hemodialysis.
‡ Greater than 200% sustained increase of creatinine from baseline.

### Secondary Outcomes

A total of 266 participants (5.5%) met the secondary composite outcome of death, renal replacement therapy, >200% sustained increase of creatinine from baseline, myocardial infarction and stroke. The care bundle strategy was associated with lower rates of renal replacement therapy (0.0% vs. 0.4%; mean difference, –0.3% [95% CI, –0.6% to –0.1%]; RR, 0.11 [95% CI, 0.01 to 0.59]; *P* = .04) and creatinine >200% of baseline (1.5% vs. 3.6%; mean difference, –2.1% [95% CI, –3.0% to –1.2%]; RR, 0.41 [95% CI, 0.28 to 0.60]; *P* < .001). The care bundle strategy was also associated with a similar risk of all-cause death, myocardial infarction, and stroke when compared with usual care.

### Subgroup and Post Hoc Analyses

In subgroup analyses, the association between care bundle strategy and lower risk of AKI remained statistically significant across all prespecified patient subgroups (Figure 1). Among eight prespecified stratified analyses for the primary outcome of AKI, there were no statistically significant interactions for all subgroups. However, the protective effects of care bundle strategy were more pronounced among female (51.0 vs. 61.8; RR, 0.83 [95% CI, 0.73 to 0.93]), patients with concomitant cardiac surgeries (54.7 vs. 65.3; RR, 0.84 [95% CI, 0.76 to 0.92]), and those undergoing CABG with cardiopulmonary bypass (55.1 vs. 62.5; RR, 0.88 [95% CI, 0.83 to 0.94]).

**Figure 1.**
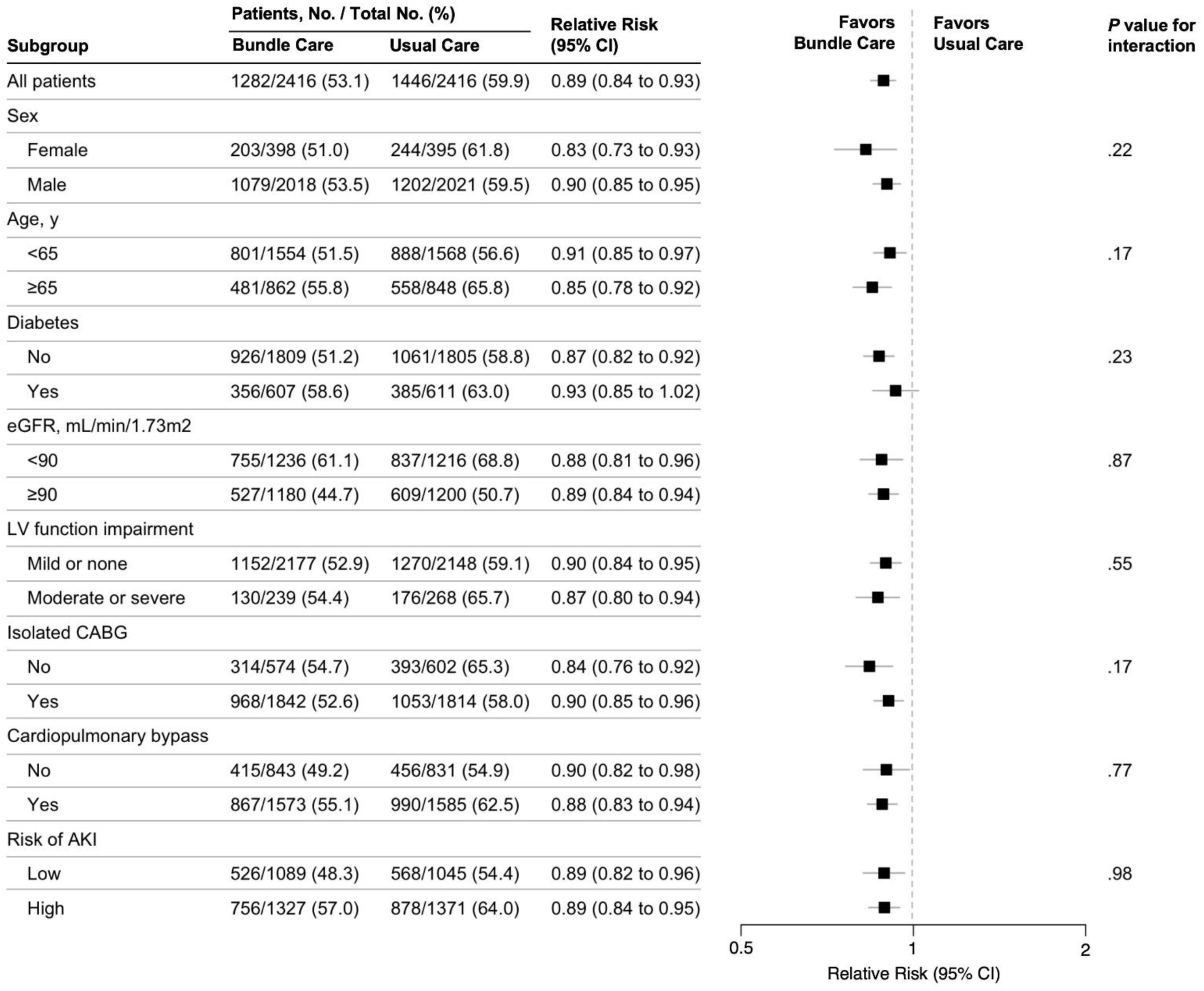
Prespecified subgroup analysis of the effect of the care bundle on acute kidney injury incidence. Relative risks (RRs) were derived from complete case data with the use of stratified generalized linear modelling with error bars indicating 95% CIs. An RR <1 indicates a decrease in AKI. CI indicates confidence interval; eGFR, estimated glomerular filtration; LV, left ventricular; AKI, acute kidney injury.

In post hoc exploratory analyses, care bundle strategy was associated with significantly lower hazards of mechanical ventilation time over 24 h (5.3% vs. 12.1%; mean difference, –6.9% [95% CI,–8.4% to –5.3%]; RR, 0.40 [95% CI, 0.32 to 0.50]; *P* < .001), intra-aortic ballon pump (0.5% vs. 2.3%; mean difference, –1.8% [95% CI, –2.4% to –1.1%]; RR, 0.23 [95% CI, 0.12 to 0.41]; *P* < .001), as well as repeat operation (0.5% vs. 1.7%; mean difference, –1.1% [95% CI, –1.7% to –0.5%]; RR, 0.33 [95% CI, 0.17 to 0.59]; *P* < .001) during the same hospitalization compared with those treated with standard care (eTable 2 in Supplement).

## Discussion

In this registry-based, propensity-matched cohort study, implementation of a prophylactic care bundle comprising evidence-based perioperative practices was associated with decreased risk of postoperative AKI compared with standard care. This protective effect was more pronounced in patients with a high risk of postoperative AKI.

Previous observational studies have reported that AKI following cardiac surgery is associated with longer hospital stays, more postoperative complications, and increased cost of care.^6,22^ Despite the clinical relevance of this condition, no pharmacological and nonpharmacological preventive strategies have been consistently proven in large phase III clinical trials to reduce the occurrence of AKI following cardiac surgery.^16,23^

The KDIGO clinical practice guideline for AKI recommends a supportive, prevention-focused bundle that includes optimization of hemodynamics, avoidance of nephrotoxins, and glycemic control.^8,9^ However, adequately powered study is warranted to investigate the effect of the care bundle approach on mortality and renal outcomes, especially following cardiac surgery. Two randomized clinical trials demonstrated that early implementation of this bundle in high-risk patients identified by postoperative biomarkers significantly reduced AKI occurrence.^11,17^ In a retrospective study, by Engelman et al. demonstrated that a multidisciplinary acute kidney response team applying bundle approach achieved an 89% relative risk reduction in stage 2 or 3 AKI after cardiac surgery.^24^ Reflecting this growing evidence the Society of Thoracic Surgeons now recommends adopting KDIGO bundle for adult patients undergoing cardiopulmonary bypass who are at high risk of AKI.^10^

Current best practice includes temporary discontinuation of ACEIs and ARBs before surgery to avoid perioperative hypotension.^9^ Although non-steroidal anti-inflammatory drugs are widely used in patients with normal kidney function, their impact on risk of AKI in the postoperative period is unclear.^25^ Intraoperative and early postoperative hemodynamic management have a major impact on the development of AKI.^26^ A higher mean arterial blood pressure target (>65 mmHg) may be beneficial in reducing the incidence of AKI in patients with poorly controlled pre-existing hypertension.^23^ As postoperative hyperglycemia is strongly associated with AKI, avoidance of perioperative hyperglycemia (>180 mg/dl) is recommended.^27^

In our study, the association between bundle care and AKI was significant among patients undergoing CABG with cardiopulmonary bypass, which is often associated with renal hypoperfusion, resulting from the low-flow, low-pressure, non-pulsatile perfusion with hemodilution and rapid temperature changes, demonstrates an increased risk of AKI.^23^ This finding aligns with meta-analytic evidence showing that minimally invasive extracorporeal circulation can reduce AKI risk by >50% compared with conventional cardiopulmonary bypass.^28^ The bundle also appeared beneficial in patients with reduced baseline renal function (eGFR <60 mL/min/1.73 m^2^), suggesting its applicability to this high-risk subgroup. However, there was no statistically significant differences in prespecified cardiovascular outcomes including myocardial infarction and stroke between the bundle and standard care groups, both in patients with high and low risks of AKI.

### Limitations

This study has certain limitations. First, as an observational study, it is subject to residual confounding and lacks independent laboratory confirmation of AKI, although falsification endpoints analyses suggested no evidence of systemic bias.

Second, data in the relative contribution of individual bundle components, such as serial serum creatinine and urine output monitoring, hemodynamic monitoring, nephrotoxin avoidance, temporary discontinuation of ACEIs and ARBs, and glycemic control, were not included. Adequately powered multicenter trials are warranted to investigate the effect of the individual approach on renal outcomes, and to explore the role of biomarkers in the identification of patients who might benefit from specific interventions.^29^

Third, outcomes beyond hospital discharge were not assessed. Severe AKI after CABF has been associated with significant risks of adverse long-term health outcomes. Considering large gaps in knowledge remain about effective interventions that can improve the outcomes of patients, novel clinical interventions are essential to identify effective therapeutic strategies and models for provision of health care that reduce the long-term sequelae of AKI.^30^

Fourth, the number of patients receiving renal replacement therapy in our study was relatively small and the confidence intervals around the point estimates were relatively wide. The initiation of dialysis was left to the judgment of the attending physician because of the lack of widely accepted criteria or guidelines for initiating renal replacement therapy. Findings for analyses of renal replacement therapy in secondary outcomes should be interpreted as exploratory.^3,5^

## Conclusions

Among patients undergoing CABG, implementation of a structured care bundle strategy during the perioperative period was associated with a lower incidence of AKI compared with standard care, especially among those at high risk of postoperative AKI. Further investigation, including from randomized clinical trials, is needed to more definitively determine the role of bundle care in patients undergoing cardiac surgery.

## Data Availability

Data of this study belongs to China National Center for Cardiovascular Diseases and will be available upon request.

## Acknowledgments

We appreciate the multiple contributions made by follow-up staffs including Li He, Yuyan Zhou, and Ying Zhang (Fuwai Hospital, National Center for Cardiovascular Diseases, Chinese Academy of Medical Sciences and Peking Union Medical College, Beijing, People’s Republic of China). No one received compensation for their role in this study.

## Nonstandard Abbreviations and Acronyms

ACE inhibitors: angiotensin-converting-enzyme inhibitors
AKI: acute kidney injury
ARB: angiotensin II receptor blockers
BMI: body mass index
CABG: coronary artery bypass grafting
CI: confidence interval
COPD: chronic obstructive pulmonary disease
eGFR: estimated glomerular filtration
EuroSCORE: European System for Cardiac Operative Risk Evaluation
IABP: intra-aortic ballon pump
ICU: intensive care unit
IQR: interquartile range
KDIGO: Kidney Disease Improving Global Outcomes
LVEF: left ventricular ejection fraction
MACKE: major adverse cardiac and kidney events
OR: odds ratio
RR: relative risk
RRT: renal replacement therapy
PCI: percutaneous coronary intervention
SD: standard deviation

## REFERENCES

1. Zarbock A, Weiss R, Albert F, Rutledge K, Kellum JA, Bellomo R, Grigoryev E, Candela-Toha AM, Demir ZA, Legros V, et al. Epidemiology of surgery associated acute kidney injury (EPIS-AKI): A prospective international observational multi-center clinical study. Intensive Care Med. 2023;49(12):1441–1455. doi: 10.1007/s00134-023-07169-7.

2. Scurt FG, Bose K, Mertens PR, Chatzikyrkou C, Herzog C. Cardiac surgery-associated acute kidney injury. Kidney360. 2024;5(6):909–926. doi: 10.34067/KID.0000000000000466.

3. Kellum JA, Prowle JR. Paradigms of acute kidney injury in the intensive care setting. Nat Rev Nephrol. 2018;14(4):217–230. doi: 10.1038/nrneph.2017.184.

4. Li S, Fu S, Xiao Y, Xu G. Recent perioperative pharmacological prevention of acute kidney injury after cardiac surgery: A narrative review. Am J Cardiovasc Drugs. 2017;17(1):17–25. doi: 10.1007/s40256-016-0194-z.

5. Ostermann M, Bellomo R, Burdmann EA, Doi K, Endre ZH, Goldstein SL, Kane-Gill SL, Liu KD, Prowle JR, Shawet AD, et al. Controversies in acute kidney injury: Conclusions from a Kidney Disease: Improving Global Outcomes (KDIGO) Conference. Kidney Int. 2020;98(2):294–309. doi: 10.1016/j.kint.2020.04.020.

6. Jacob KA, Leaf DE. Prevention of cardiac surgery-associated acute kidney injury: A review of current strategies. Anesthesiol Clin. 2019;37(4):729–749. doi: 10.1016/j.anclin.2019.08.007.

7. Hariri G, Collet L, Duarte L, Martin GL, Resche-Rigon M, Lebreton G, Bouglé A, Dechartres A. Prevention of cardiac surgery-associated acute kidney injury: A systematic review and meta-analysis of non-pharmacological interventions. Crit Care. 2023;27(1):354. doi: 10.1186/s13054-023-04640-1.

8. Khwaja A. KDIGO clinical practice guidelines for acute kidney injury. Nephron Clin Pract. 2012;120(4):c179–c184. doi: 10.1159/000339789.

9. Joannidis M, Druml W, Forni LG, Groeneveld ABJ, Honore PM, Hoste E, Ostermann M, Straaten HMO, Schetz M. Prevention of acute kidney injury and protection of renal function in the intensive care unit: Update 2017: Expert opinion of the Working Group on Prevention, AKI section, European Society of Intensive Care Medicine. Intensive Care Med. 2017;43(6):730–749. doi: 10.1007/s00134-017-4832-y.

10. Brown JR, Baker RA, Shore-Lesserson L, Fox AA, Mongero LB, Lobdell KW, LeMaire SA, Somer FMJJD, Ballmoos MW, Barodka V, et al. The Society of Thoracic Surgeons/Society of Cardiovascular Anesthesiologists/American Society of Extracorporeal Technology Clinical Practice guidelines for the prevention of adult cardiac surgery-associated acute kidney injury. Ann Thorac Surg. 2023;115(1):34–42. doi: 10.1016/j.athoracsur.2022.06.054.

11. Göcze I, Jauch D, Götz M, Kennedy P, Jung B, Zeman F, Gnewuch C, Graf BM, Gnann W, Banas B, et al. Biomarker-guided intervention to prevent acute kidney injury after major surgery: The prospective randomized BigpAK study. Ann Surg. 2018;267(6):1013–1020. doi: 10.1097/SLA.0000000000002485.

12. James MT, Har BJ, Tyrrell BD, Faris PD, Tan Z, Spertus JA, Wilton SB, Ghali WA, Knudtson ML, Sajobi TT, et al. Effect of clinical decision support with audit and feedback on prevention of acute kidney injury in patients undergoing coronary angiography: A randomized clinical trial. JAMA. 2022;328(9):839–849. doi: 10.1001/jama.2022.13382.

13. Wilson FP, Yamamoto Y, Martin M, Coronel-Moreno C, Li F, Cheng C, Aklilu A, Ghazi L, Greenberg JH, Latham S, et al. A randomized clinical trial assessing the effect of automated medication-targeted alerts on acute kidney injury outcomes. Nat Commun. 2023;14(1):2826. doi: 10.1038/s41467-023-38532-3.

14. Rao C, Zhang H, Gao H, Zhao Y, Yuan X, Hua K, Hu S, Zheng Z; Chinese Cardiac Surgery Registry Collaborative Group. The Chinese cardiac surgery registry: Design and data audit. Ann Thorac Surg. 2016;101(4):1514–1520. doi: 10.1016/j.athoracsur.2015.09.038.

15. D’Agostino RS, Jacobs JP, Badhwar V, Paone G, Rankin JS, Han JM, McDonald D, Shahian DM. The Society of Thoracic Surgeons adult cardiac surgery database: 2016 update on outcomes and quality. Ann Thorac Surg. 2016;101(1):24–32. doi: 10.1016/j.athoracsur.2015.11.032.

16. Thielmann M, Corteville D, Szabo G, Swaminathan M, Lamy A, Lehner LJ, Brown CD, Noiseux N, Atta MG, Squiers EC, et al. Teprasiran, a small interfering RNA, for the prevention of acute kidney injury in high-risk patients undergoing cardiac surgery: A randomized clinical study. Circulation. 2021;144(14):1133–1144. doi: 10.1161/CIRCULATIONAHA.120.053029.

17. Meersch M, Schmidt C, Hoffmeier A, Aken HV, Wempe C, Gerss J, Zarbock A. Prevention of cardiac surgery-associated AKI by implementing the KDIGO guidelines in high risk patients identified by biomarkers: The PrevAKI randomized controlled trial. Intensive Care Med. 2017;43(11):1551–1561. doi: 10.1007/s00134-016-4670-3.

18. Maeda A, Inokuchi R, Bellomo R, Doi K. Heterogeneity in the definition of major adverse kidney events: A scoping review. Intensive Care Med. 2024;50(7):1049–1063. doi: 10.1007/s00134-024-07480-x.

19. Dehmer GJ, Badhwar V, Bermudez EA, Cleveland JC, Cohen MG, D’Agostino RS, Ferguson TB, Hendel RC, Isler ML, Jacobs JP, et al. 2020 AHA/ACC Key data elements and definitions for coronary revascularization: A report of the American College of Cardiology/American Heart Association task force on clinical data standards (writing committee to develop clinical data standards for coronary revascularization). J Am Coll Cardiol. 2020;75(16):1975–2088. doi: 10.1016/j.jacc.2020.02.010.

20. Austin PC. Optimal caliper widths for propensity-score matching when estimating differences in means and differences in proportions in observational studies. Pharm Stat. 2011;10(2):150–161. doi: 10.1002/pst.433.

21. Austin PC. Balance diagnostics for comparing the distribution of baseline covariates between treatment groups in propensity-score matched samples. Stat Med. 2009;28(25):3083–3107. doi: 10.1002/sim.3697.

22. Forni LG, Darmon M, Ostermann M, Straaten HMO, Pettilä V, Prowle JR, Schetz M, Joannidis M. Renal recovery after acute kidney injury. Intensive Care Med. 2017;43(6):855–866. doi: 10.1007/s00134-017-4809-x.

23. Wang Y, Bellomo R. Cardiac surgery-associated acute kidney injury: Risk factors, pathophysiology and treatment. Nat Rev Nephrol. 2017;13(11):697–711. doi: 10.1038/nrneph.2017.119.

24. Engelman DT, Crisafi C, Germain M, Greco B, Nathanson BH, Engelman RM, Schwann TA. Using urinary biomarkers to reduce acute kidney injury following cardiac surgery. J Thorac Cardiovasc Surg. 2020;160(5):1235–1246.e2. doi: 10.1016/j.jtcvs.2019.10.034.

25. Bell S, Rennie T, Marwick CA, Davey P. Effects of peri-operative nonsteroidal anti-inflammatory drugs on post-operative kidney function for adults with normal kidney function. Cochrane Database Syst Rev. 2018;11(11):CD011274. doi: 10.1002/14651858.CD011274.pub2.

26. Jentzer JC, Bihorac A, Brusca SB, Rio-Pertuz GD, Kashani K, Kazory A, Kellum JA, Mao M, Moriyama B, Morrow DA, et al. Contemporary management of severe acute kidney injury and refractory cardiorenal syndrome: JACC council perspectives. J Am Coll Cardiol. 2020;76(9):1084–1101. doi: 10.1016/j.jacc.2020.06.070.

27. Frisch A, Chandra P, Smiley D, Peng L, Rizzo M, Gatcliffe C, Hudson M, Mendoza J, Johnson R, Lin E, et al. Prevalence and clinical outcome of hyperglycemia in the perioperative period in noncardiac surgery. Diabetes Care. 2010;33(8):1783–1788. doi: 10.2337/dc10-0304.

28. Kowalewski M, Pawliszak W, Raffa GM, Malvindi PG, Kowalkowska ME, Zaborowska K, Kowalewski J, Tarelli G, Taggart DP, Anisimowicz L. Safety and efficacy of miniaturized extracorporeal circulation when compared with off-pump and conventional coronary artery bypass grafting: Evidence synthesis from a comprehensive Bayesian-framework network meta-analysis of 134 randomized controlled trials involving 22 778 patients. Eur J Cardiothorac Surg. 2016;49(5):1428–1440. doi: 10.1093/ejcts/ezv387.

29. Demirjian S, Bashour CA, Shaw A, Schold JD, Simon J, Anthony D, Soltesz E, Gadegbeku CA. Predictive accuracy of a perioperative laboratory test-based prediction model for moderate to severe acute kidney injury after cardiac surgery. JAMA. 2022;327(10):956–964. doi: 10.1001/jama.2022.1751.

30. James MT, Bhatt M, Pannu N, Tonelli M. Long-term outcomes of acute kidney injury and strategies for improved care. Nat Rev Nephrol. 2020;16(4):193–205. doi: 10.1038/s41581-019-0247-z.

